# Thrombotic microvascular injury is not mediated by thrombotic microangiopathy despite systemic complement activation in Covid-19 patients

**DOI:** 10.1101/2020.06.18.20115873

**Authors:** Adrien De Voeght, Doriane Calmes, Floran Beck, Jean-Baptiste Sylvestre, Philippe Delvenne, Pierre Peters, Gaelle Vertenoeil, Frédéric Baron, Nathalie Layios, Jean-Luc Canivet

## Abstract

Hypoxemia and coagulopathy are common in severe symptomatic patients of coronavirus disease 2019 (COVID-19). Histological evidence shows implication of complement activation and lung injury. We research sign of complement activation and presence of thrombotic microangiopathy in 8 severe patients. Six of them presented moderate elevation of final pathway of complement – sC5b-9 (median value : 350 ng/mL [IQR : 300,5 - 514,95 ng/mL]). Two patients have been autopsied and presence of thrombotic microvascular injury have been found. Interestingly, none the 8 patients had signs of mechanical hemolytic anemia (median value of hemoglobin : 10,5 gr/dL[IQR : 8,1 - 11,9], median value of haptoglobuline 4,49 [IQR 3,55-4,66], none of the patients has schistocyte) and thrombocytopenia (median value: 348000/mL [IQR : 266 000 - 401 000). Finally, all 8 patients had elevated d-dimer (median value : 2226 µgr/l [IQR : 1493 – 2362]) and soluble fibrin monomer complex (median value : 8.5 mg/mL, IQR[<6 – 10.6]). In summary, this study show moderate activation of complement and coagulation with presence of thrombotic microvascular injury in patients with severe COVID-19 without evidence of systemic thrombotic microangiopathy.

**Key points:**

1. Severe covid-19 patients show moderate elevation of final activation of complement
2. No sign of Thrombotic microangiopathy is found in severe covid-19 patients

## Introduction

In March 2020, COVID-19 was described as a pandemic by the World Health Organisation (1). Mortality and intensive care requirement are high among patients with respiratory failure and severe hypoxemia (2).

Early reports of autopsies showed classical lesions of acute respiratory distress syndrome (ARDS) such as diffuse alveolar damage and hyaline membrane formation. Recent reports display diffuse injuries of alveolar capillaries which might be related to thrombotic microvascular injury rather than classical image of ARDS (3,4).These microvascular injuries appear to occur early in the pathogenicity of the disease and could be linked to ACE2 activity dysregulation leading to lung blood flow redistribution as seen in an experimental rat model (5). This could partially explain why severe hypoxemia coexists with preserved lung compliance, hence increased shunt effect and high dead space (6).

Thrombotic microangiopathy (TMA) is suspected to play a role in the physiopathology of Covid-19 hypoxemia without convincing biological data (7). TMA is a complex entity associated with occlusion of small and large vessels, which encompasses pathologies such as thrombotic thrombocytopenic purpura, hemolytic uremic syndrome (HUS) and other TMA related to medication, malignancies or infection (8). All of these entities have in common a mechanic hemolytic anemia (HA) associated with thrombocytopenia. Atypical TMA (aHUS) are associated either with dysregulation of complement activation or endothelial injury or both. Evidences from SARS-CoV and MERS-CoV show an association between complement activation and severity of these diseases. The same has been shown in an animal Covid-19 model, in which it was demonstrated that N proteins of SARC-CoV2 bind to mannose binding lectin (MBL) -associated serine protease 2 (MASP-2), the key serine protease in the lectin pathway of complement and that lead to complement hyperactivation. It was also demonstrated a correlation between complement hyperactivation and severity of lung injury (9-11). Furthermore, autopsy data show co-localization of viral spike glycoprotein and components of lectin and alternative pathway complement in the lungs of patients with covid-19 pneumonia. This was associated with microthrombosis (4) and supported hypothesis of TMA (7) occurring in Covid-19 patients.

Complement system is a part of innate immunity. Three pathways (classical by interaction between antigens and antibody complexes, alternative by stimulation of specific antigen and lectinby binding mannose residue on the pathogen surface) lead to a final activation with a lytic formation (membrane attack complex - MAC) on the cellular surface resulting in destruction of this cell (12).

We aimed to explore whether Covid-19 severely ill patients had systemic complement activation and whether it was potentially attributable to TMA, in order to explain resulting microvascular thrombosis in the lung.

## Methods

We performed observational study in eight critically ill patients with severe COVID-19 pneumonia (mean age of 63 years (SD : 61-67 year), mean SAPS2 of 39 (SD : 34-43) and mean PaFi 190 (SD : 126-243). We collected biological markers of TMA, thrombotic state (D-Dimer and soluble fibrin monomer complex (FMC)) and complement activation. All the patients were mechanically ventilated.

### Complement analyses

Serum was collected after centrifugation. Serum levels of C3c and C4 was measured by nephelometry (Siemens Healthcare, Marburg, Germany). CH50 was measured by turbidimetrics assays (Optilite System, Binding Site, Birmingham, UK). SC5b-9 (MAC) was assessed by Microvue SC5b-9 plus ELISA Kit (Quidek Co., San Diego, USA).

### Systemic TMA

TMA is defined as the association of mechanical hemolytic anemia (MAHA) (LDH, acute anemia and schistocytes above 2%) and thrombocytopenia (13). Schistocytes were characterized and counted according to ICSH recommendations (14).

### Autopsy

Autopsies were undertaken following recent Covid-19 guidelines (15).Lung specimens were examined by direct and microscopic exam including specific coloration such as immunohistochemistry.

### Statistical analyses

Continuous variables are presented as median and interquartile range values (IQR); categorical variables are expressed as number (%).

## Results and discussion

None of the eight patients met the criteria of TMA (Table 1) 0/7 patients had schistocytes above 1%. Anemia (median value: 10,5 gr/dL[IQR : 8,1 - 11,9]) was deemed to be related to hyperinflammation in all cases. LDH was elevated (median value: 504 U/L [IQR : 324 −754]). No thrombocytopenia was observed (median value: 348000/mL [IQR : 266 000 - 401 000]).

**Table 1:**
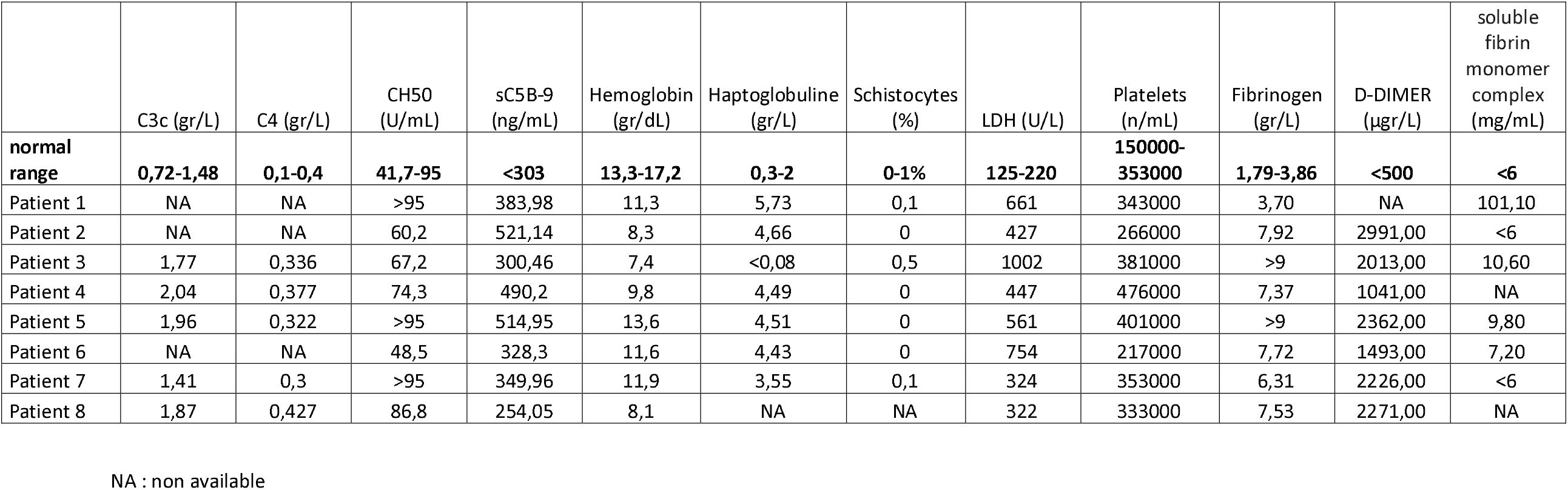
biological parameters.

Exploration of terminal activation of complement showed moderate activation for 6/8 of the patients (median value : 350 ng/mL [IQR : 300,5 - 514,95 ng/mL]) CH50 level was elevated in 3/8 patients (median value : 80.5 U/mL [IQR : 60.2 - >95]). C3c levels were moderate elevated in 4/5 patients (median value :1,87 gr/L [IQR :1,77 - 1,96]), C4 level is high in only 1/5 patients (median value : 0,336 gr/L [IQR : 0,322-0,377]).

Fibrinogen was high in 7/8 patients (median value : 7.63 [IQR 6.31-9]). D-Dimers were markedly increased in 7/7 patients (median value : 2226 µgr/l [IQR : 1493 – 2362]). FMC were high in 4/6 patients (median value : 8.5 mg/mL, IQR[<6 – 10.6]).

50% of patients died. Two autopsies were performed (patients 1 and 6). Disseminated microvascular thrombosis was found in the lungs (Figure 1). In contrast, kidneys were not affected. These two patients had signs of systemic complement and coagulation activation (high levels of FMC and D-dimers).

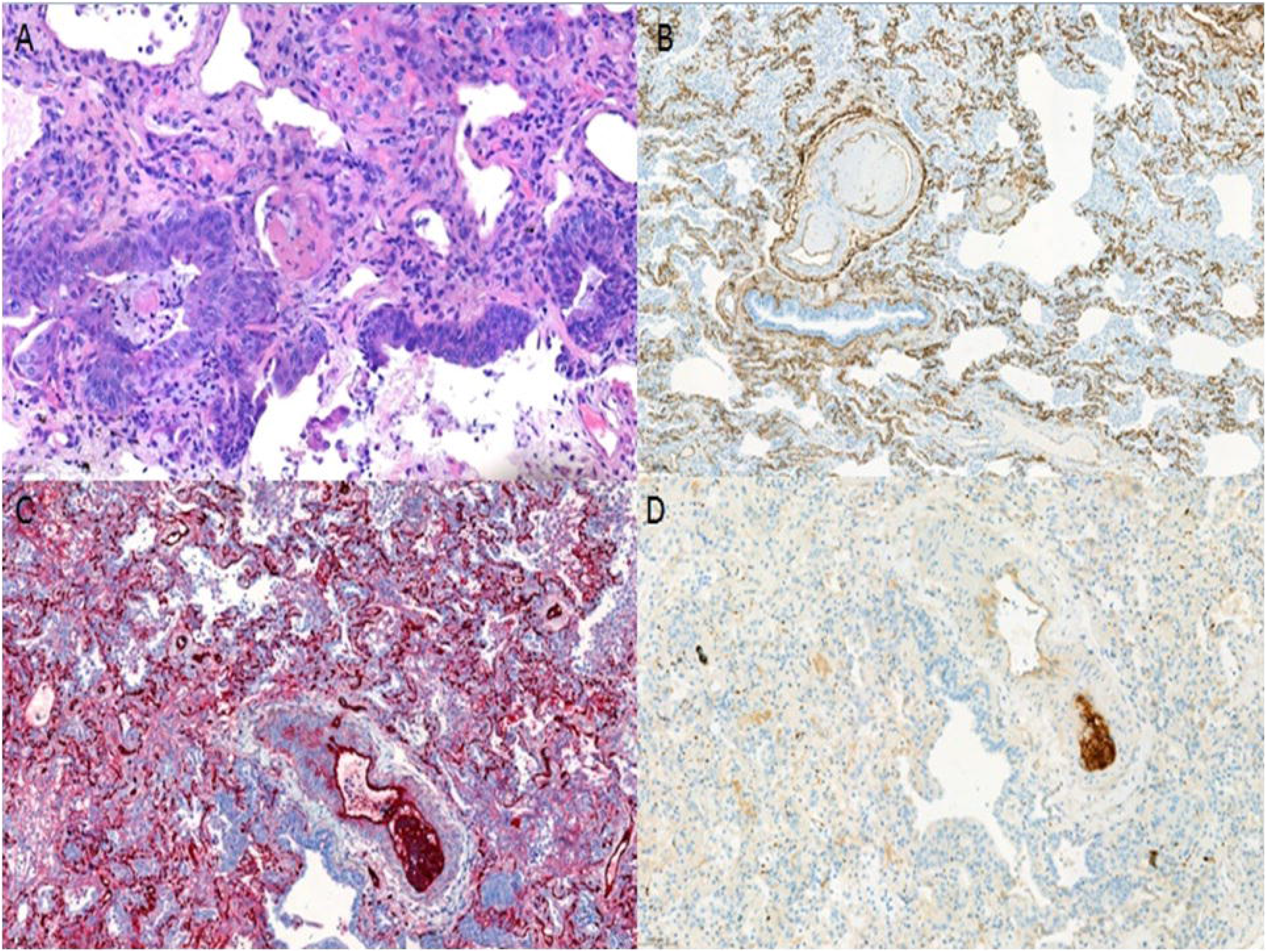
A-D. Demonstration of partially occluded microvascular structures in lung tissue specimens from patient 6.A. Hematoxylin eosin. B-D. Immunohistology with antibodies against CD34 (B), FVIII (C) and CD61 (D)

Our data show moderate activation of the terminal complement system in 75% of our patients. Elevated markers of direct pathway are probably linked mainly to systemic observation. Evidence for a prothrombotic state was found with elevated levels of D-Dimers and FMC.

This observation confirms previous post-mortem findings showing microthrombosis inside lung vessels (4). Nevertheless, we do not find any presence of MAHA and thrombocytopenia which is the definition of systemic TMA (13).

A previous autopsy study found important deposits of C5b-9 in lung vessels, C4d in inter-alveolar septa around microvascular injury and MBL-MASP-2 around inter-alveolar septa suggesting an activation of both alternative and lectine complement pathways (4). Recent experimental data demonstated that N-protein of COVID-19 can bin to MASP2 which lead to aberrant activation of complement and are associated whith severity of lung injury. Moreover, this work has showed that specific blocking of lectin pathway lead to decrease in lung injury (11).

Recently, it has been suggested that TMA could be the link between SARS-CoV2 and the thrombotic microvascular lung injury observed in autopsy studies (7). Our data do not support this hypothesis. An alternative explanation could be that local activation of the complement is triggered by direct viral injury resulting in endothelial damage and vascular microthrombosis in the absence of any systemic TMA. Indeed, coronaviruses activate innate immunity and increase complement activation without any TMA (11). This local activation of complement could result in endothelial injury (16). This could induce coagulation activation (17). Complement dysregulation can directly lead to activation of coagulation resulting in microthrombosis(18, 19).

Taken together, these data might support the rational use of anti-complement drugs such as anti-C5 monoclonal antibodies (eculizumab, ravulizumab), which are currently evaluated in multiple ongoing phase III trials (20). Alternatively, anti-C3 might also be considered as recently suggested (21).

In summary, thrombotic microvascular injury observed in the lung of critically ill patient with severe covid-19 pneumonia is associated with systemic activation of coagulation (as evidenced by D-Dimers and FMC elevation) and complement activation without evidence of TMA. Association between complement activation secondary to endothelial injury and prothrombotic coagulopathy seems to be one of the key pathological features of Covid-19 pneumonia.

## Data Availability

Data is available

## Authorship contribution

ADV, NL and JLC designed the study; ADV, DC, FB, JBS collected biological and clinical data; ADV, FB, PP, GV, NL and JLC interpreted clinical data; ADV and PD interpreted pathology data; ADV, FB and JLC drafted the manuscript; and all authors approved the final manuscript

## Disclosure of conflicts of interest

The authors declare no competing financial interests

## Notes

### Competing Interest Statement

The authors have declared no competing interest.

### Funding Statement

no grant hase been received

### Author Declarations

all ethical guidelines have been followed The protocol was approved by the Ethics Committee of the University Hospital of Liege (2020/214)

